# Agentic AI versus pediatric emergency physicians for differential diagnosis of acutely ill and injured children

**DOI:** 10.64898/2026.11.16.26367440

**Authors:** Nathan Georgette, Thomas A Buckley, Andrew M. Fine, James Jones, Andrew J. McMurry, Alexandra T. Geanacopoulos, Katia C. Genadry, Robert Hoffmann, Caroline G. Kahane, Arjun K Manrai, Kenneth D Mandl

**Author notes:** NG and TAB share first authorship of this study. AKM and KDM share senior authorship of this study. Corresponding author: Kenneth D. Mandl, MD, MPH, Director, Computational Health Informatics Program, Boston Children’s Hospital, 401 Park Drive, Landmark 5506, Mail Stop BCH3187, Boston, Massachusetts 02215. **Contributors** KDM and AKM conceptualized the study and were responsible for overall supervision. NG, TAB, AMF, KDM, and AKM designed the study. JJ and AJMcM developed and implemented code for case selection and text processing. TAB and AKM developed and implemented the Dr. CaBot AI agent. NG, AMF, ATG, KCG, RH, CGK performed case reviews and assessments. NG, TAB, and AMF had access to and verified all underlying data. NG performed the statistical analyses and wrote the original manuscript draft. All authors reviewed and edited the manuscript, had full access to the study data, and had final responsibility for the decision to submit for publication.

## Abstract

**Background:** Misdiagnosis and delayed diagnosis of children in pediatric emergency departments lead to complications, and are particularly problematic in settings with limited pediatric expertise. Large language models (LLMs) have shown promise in diagnostic reasoning for adult emergency department patients, but utility for pediatric cases is largely untested.

**Methods:** Retrospective study of patients seen in the emergency department of a major urban pediatric academic medical center and subsequently admitted to the operating room or inpatient service. Records from 100 patients were randomly selected, stratified by age. We compared the accuracy of differential diagnoses (up to five, ranked by likelihood) generated by a previously developed agentic AI system, Dr. CaBot (one configuration with literature search, the other without), with those of four attending pediatric emergency medicine physicians (diagnosticians), all masked to outcome. Encounters were presented to diagnosticians stepwise across four touchpoints: triage, the initial physician evaluation (the *a priori* primary touchpoint), the full emergency department encounter, and the admission. Ground truth diagnosis was determined by two independent, board-certified pediatric emergency physicians (reviewers) based on the entire clinical encounter. The reviewers, masked to source, then assigned each differential a Bond score (0 to 5; 4 or above was high-scoring).

**Findings:** The median patient age was 4 years and 41% had a complex chronic condition. Bond score agreement between the two physician reviewers had a weighted kappa of 0.64. For the primary comparison at the initial physician evaluation touchpoint, Dr. CaBot and the diagnosticians both achieved high-scoring differentials in 95% of cases (difference 0%, 95%CI: -5.8, 5.8%). No differences were detected at the other touchpoints.

**Interpretation:** Dr. CaBot and attending pediatric emergency medicine physicians showed similar performance in generating differential diagnoses from clinical documentation, suggesting potential as a diagnostic aid. Whether this performance can be translated into improvements in care requires prospective evaluation.

**Research in context:** *Evidence before this study:* We searched PubMed using the query ((Pediatric[Title/Abstract]) AND (Emergency medicine[Title/Abstract]) AND ((Artificial Intelligence[Title/Abstract]) OR (Diagnostic reasoning[Title/Abstract]))), for articles published from 1 January 2000 to 31 December 2025, yielding 28 results, of which 16 were deemed not relevant based on title, 6 were reviews not presenting novel data, and 3 were surveys of physician perspectives on artificial intelligence (AI). There were 3 remaining studies of AI applications to pediatric emergency populations. One assessed AI management decisions of critically ill patients. One attempted to automate triage categorization using machine learning. One assessed the diagnostic reasoning of various general large language models (LLMs; e.g. ChatGPT-4o) on adapted cases from a pediatric emergency department in Italy. Overall, there is limited data on the use of specialized AI agents on diagnostic reasoning on real-world pediatric emergency cases.

*Added value of this study:* The diagnostic accuracy of an LLM-based agentic process was similar to that of four attending pediatric emergency medicine physicians across 100 cases of children evaluated in a pediatric emergency department. Using real world data from electronic health records and a masked multi-physician evaluation of open-ended agentic responses, we demonstrated physician-level accuracy of artificial intelligence in generating differential diagnoses for children.

*Implications of all the available evidence:* The agent shows promise as a diagnostic aid among children presenting to the emergency department.

## Background

Artificial intelligence (AI) has demonstrated capacity for accurate diagnosis including robust clinical reasoning through complex cases.^1–8^ A retrospective cohort study of adults presenting to the emergency department (ED) found that OpenAI’s o1 model could generate differential diagnoses with accuracy on par with board-certified internists overall and with greater accuracy at the first touchpoint^9^. More recently, agentic systems have been built on these models specifically for diagnostic reasoning. One such system, Dr. CaBot, iteratively searches the medical literature and produces verifiable clinical reasoning;^8^ its differential diagnosis has been published alongside an expert discussant’s in the *New England Journal of Medicine*.^7^

Children present distinct diagnostic challenges, and misdiagnosis leads to serious complications^10^ The history is often obtained from surrogates such as the parent or other caretaker, not the patients themselves. Pre-test probability of disease varies by age. The differential diagnosis for vomiting in a neonate (e.g., volvulus, pyloric stenosis, reflux, etc.) differs from that in a teenager (e.g., appendicitis, gastroenteritis, cannabis hyperemesis, etc.). Normal ranges for vital signs vary by age and are imprecise.^11^ The wide range of congenital conditions adds further complexity to the diagnosis of the acutely ill child. Further, general and community emergency departments, where more than 67% of US children have timely access to emergency care,^12^ have higher rates of inaccurate triage than centers with pediatric specialization.^13^

To date, evaluations of the diagnostic and management reasoning abilities of large language models (LLMs) have focused overwhelmingly on adult patients. There remains limited study of AI using data from real pediatric emergency patients, with the few pediatric studies published examining AI’s clinical decisions for critically ill patients^14^ and automating triage.^15^ One study used cases adapted from real-world emergency department patients to assess the diagnostic reasoning of general LLMs, such as ChatGPT-4o, and found they had similar accuracy scores to pediatric physicians.^16^ Notably, this study used vignettes specifically designed for LLM processing, rather than raw clinical notes.

We sought to assess differential diagnoses generated by a previously developed agentic AI system (Dr. CaBot) for children presenting to a pediatric ED. We hypothesized that Dr. CaBot would achieve similar diagnostic accuracy to that of pediatric emergency physicians.

## Methods

### Study Design/Setting/Subjects

This is a retrospective study of patients seen in the emergency department of Boston Children’s Hospital, a major urban pediatric academic medical center, and subsequently admitted to the operating room or inpatient service. Cases included had an ED encounter followed by an inpatient encounter or an operative note dated within 24 hours.

We randomly selected 25 age-stratified qualifying encounters, occurring July 1, 2024 through June 30 2025, from each of four age bands: under 1 year; 1 to under 5 years; 5 to under 12 years; 12 to under 18 years. Transfers into the ED were included. Encounters were excluded if the patient was primarily cared for by one of the study authors during that encounter.

### Data

Electronic health record data was parsed into four touchpoints. (1) Triage, including the nurse triage notes and vital signs; (2) Initial physician evaluation including vital signs, the history of present illness, review of systems, past medical and surgical histories, allergies, and physical exam as documented by the treating provider; (3) Emergency department stay, with medical decision making, diagnostic reasoning, management plan, and standard data incorporated into the note such as vital signs, laboratory results, medications, radiology reports, and other interventions; (4) Admission (or operating room) history & physical, capturing the admitting (or operating) team’s full documented history and physical as well as assessment and plan.

### LLM Agent Assessment

The overall process flowchart is presented graphically in **Figure 1**.

**Figure 1.**
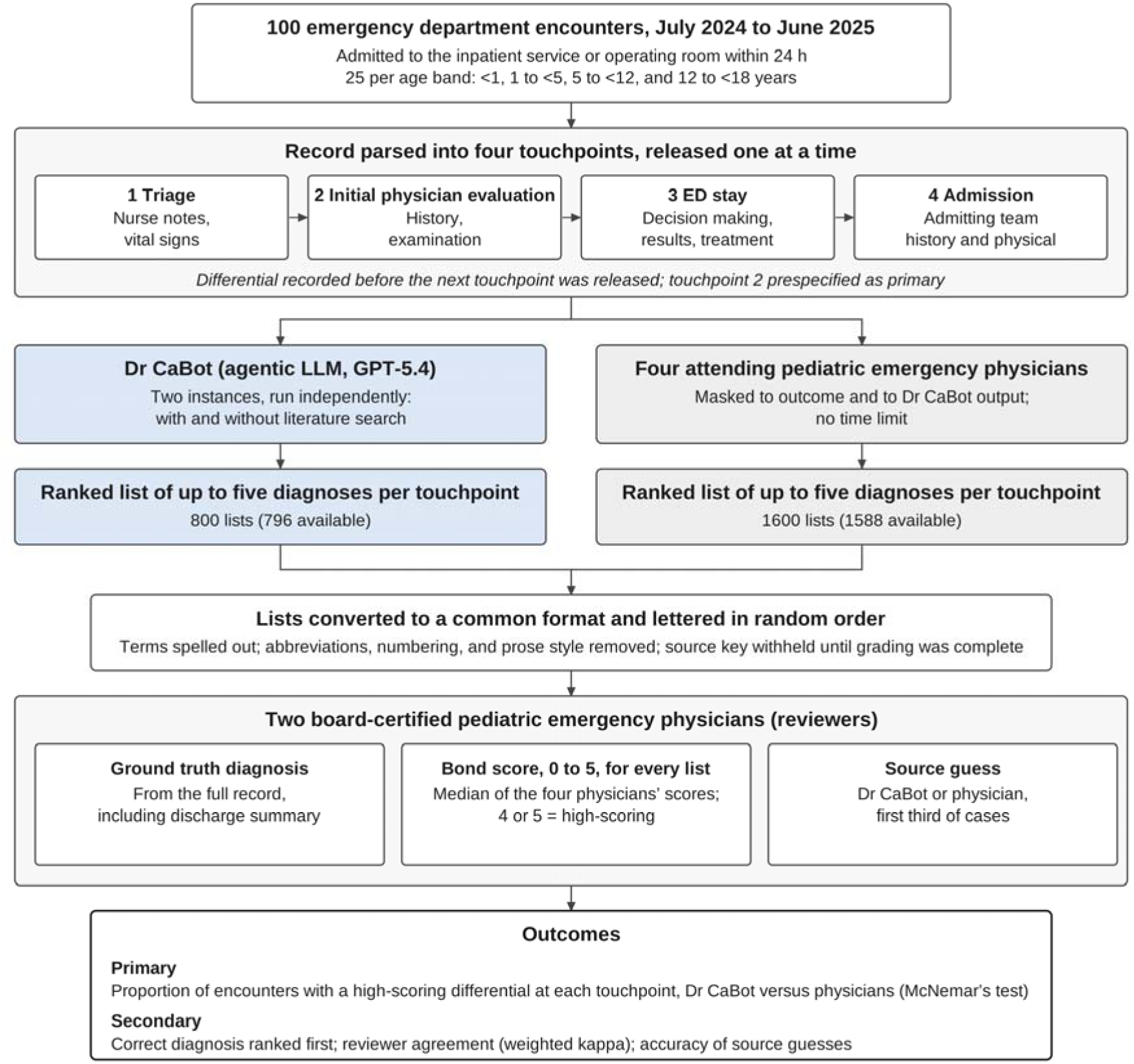
Study design and flow.

Dr. CaBot is an agentic AI system that generates a differential diagnosis by iteratively searching the medical literature and reasoning in the style of a clinicopathological conference.^8^ The prompt was adapted for the pediatric setting: the model was cast as a physician practicing in the pediatric emergency department of the study institution. Two configurations of Dr. CaBot were used independently: one was provided search access to 1.6 million clinical abstracts, as in prior work; the other was not. The search-enabled instance was allowed up to 25 calls to the literature search tool. Its base model was GPT-5.4 (OpenAI) with reasoning effort set to "high". All model queries were executed on the hospital’s high-performance computing cluster and routed exclusively to a HIPAA-compliant Azure-hosted endpoint of GPT-5.4 operated under the institution’s enterprise agreement; no patient data was sent to public OpenAI endpoints.

Data from each touchpoint was provided sequentially to Dr. CaBot. The same data were provided, in the same sequence, to four board-certified or -eligible pediatric emergency medicine attending physicians (diagnosticians), who did not see the Dr. CaBot output. The diagnosticians had no time limit. Each differential was recorded before information from the subsequent touchpoint was provided. Each diagnostician and Dr. CaBot generated a list of up to five diagnoses, ranked by likelihood.

To establish ground truth diagnosis, two board-certified pediatric emergency medicine physicians (reviewers) analyzed the patient’s medical record from the encounter, including the discharge summary, to determine the primary diagnosis. Disagreements were resolved by discussion.

To mask the reviewers to the source of each differential, diagnoses from Dr. CaBot and the diagnosticians were converted to a common format before grading, following the approach used in a prior study of adult emergency department patients^9^. Only Dr. CaBot’s final ranked list was retained; its full-text reasoning was omitted. All lists, from both sources, were expanded to fully spelled-out terms with a fixed prompt to the same GPT-5.4 deployment (e.g., "PE" to "pulmonary embolism"), removing differences in abbreviation, numbering, and prose style that could reveal the source. Within each case and touchpoint, the resulting differentials were presented to the reviewers in random order.

For the first third of cases, the reviewers were also asked to guess whether each list came from Dr. CaBot or a diagnostician. The accuracy of those guesses was calculated.

Finally, the two reviewers individually graded the accuracy of the differentials using Bond scores (**Table 1**).^17^ Therefore, each differential received two Bond scores. Bond scores are an established measure of the accuracy and usefulness of differential diagnoses generated by computer programs and AI agents.^9,17^

**Table 1.** Bond score scale^17^.

| **Bond Score\*** | **Description** |
|---|---|
| 5 | The actual diagnosis was suggested in the differential |
| 4 | The suggestions included something very close, but not exact |
| 3 | The suggestions included something closely related that might have been helpful |
| 2 | The suggestions included something related, but unlikely to be helpful |
| 0 | No suggestions close to the target diagnosis |

\*the scale does not include a score of 1

### Statistical Analysis

For each case and touchpoint, we separately calculated the median of the four diagnosticians’ and the two Dr. CaBot configurations’ Bond scores. A median Bond score of 4 or above was defined as high-scoring. For each touchpoint, the proportion of high-scoring differentials from Dr. CaBot was compared with that of the diagnosticians across the 100 encounters. Touchpoint 2 (“initial physician evaluation”) was *a priori* defined as the primary comparator due to the undifferentiated nature of the patient at that touchpoint. McNemar’s test was applied to test the null hypothesis that the paired proportions were equal. *P*-values less than 0.05 were considered statistically significant; no adjustment for multiple comparisons was made.

In an exploratory analysis of rank, we determined the position of the correct diagnosis, if present, in each differential. A differential was deemed correct if the median position was first. A sensitivity analysis excluding patients transferred from another acute care hospital (n=10) was also performed because patients transferred in may have had pre-existing diagnostic data or impressions.

The sample size was calculated based on the analogous adult emergency department study,^9^ which found a difference of 0.1 in the proportion of high-scoring differentials. For 80% power at alpha 0.05, McNemar’s test required a minimum of 77 encounters.

REDCap^18,19^ hosted at Boston Children’s Hospital was used for electronic data capture. Statistical analyses were performed using Stata 19 (StataCorp, College Station, TX). The manuscript was prepared in concordance with the STARD-AI guidelines.^20^ This study was approved by the Boston Children’s Hospital Institutional Review Board, which granted a waiver of informed consent.

## Results

### Patient characteristics

The median age was 4.0 (IQR 0.7 – 11.0) years, 45% were female, and 41% had at least one complex chronic condition.^21^ The most common chief complaints were fever (17%), respiratory symptoms (cough, shortness of breath, or wheeze; 16%), and abdominal pain (9%) (**Table 2**). The ground truth diagnoses were diverse and are presented in **Table S1**.

**Table 2.** Sample characteristics (N=100)

| **Characteristic** | **Value** |
|---|---|
| Age, yrs (IQR) | 4.0 (0.7 – 11.0) |
| Female sex (%) | 45 (45%) |
| Transfer-in from another acute care hospital | 10 (10%) |
| Complex Chronic Condition | 41 (41%) |
| Chief complaint |  |
| Fever | 17 (17%) |
| Respiratory | 16 (16%) |
| Abdominal Pain | 9 (9%) |
| Neuro/seizure | 9 (9%) |
| Vomiting | 8 (8%) |
| Others | 41 (41%) |
| Season at ED check-in |  |
| Spring | 22 (22%) |
| Summer | 14 (14%) |
| Autumn | 32 (32%) |
| Winter | 32 (32%) |

### Differentials

Across the 100 cases, the two instances of Dr. CaBot generated 796 (4 unavailable) differentials and the four diagnosticians generated 1,588 (12 unavailable). Unavailable differentials (<0.7%) were due to inadvertently skipped or missed touchpoints. Every touchpoint had at least one Dr. CaBot differential and at least two diagnostician differentials.

For 34 cases, both masked reviewers guessed the source (Dr. CaBot vs diagnostician) of the differentials, yielding 808 inferences from each reviewer. Combined, the reviewers correctly identified the source 91.6% of the time, higher than expected from chance alone (*P* < 0.001). The reviewers felt that the Dr. CaBot differentials were likely to be more verbose, esoteric, and/or comprehensive. The two reviewers showed substantial agreement in Bond scores, with a weighted kappa of 0.64.

### Bond scores

The diagnosticians’ median Bond score was high-scoring in 91, 95, 96, and 98 cases for touchpoints 1, 2, 3, and 4, respectively. For Dr. CaBot, the median Bond score was high-scoring in 91, 95, 98, and 99 cases (**Figure 2**). For the primary comparison at the initial physician evaluation touchpoint, Dr. CaBot and the diagnosticians both achieved high-scoring differentials in 95% of cases (difference 0%, 95%CI: -5.8, 5.8%). There was no difference between the diagnosticians and Dr. CaBot in these pairwise comparisons (*P* > 0.3 at each touchpoint). We found that the addition of clinical abstracts did not detectably improve the diagnostic accuracy of Dr. CaBot in this setting (**Table S2**). The proportions of high-scoring differentials for each diagnostician are shown in **Table S3**. A sensitivity analysis excluding transfers in (N=90) had unchanged overall results (**Figure S1**).

**Figure 2.**
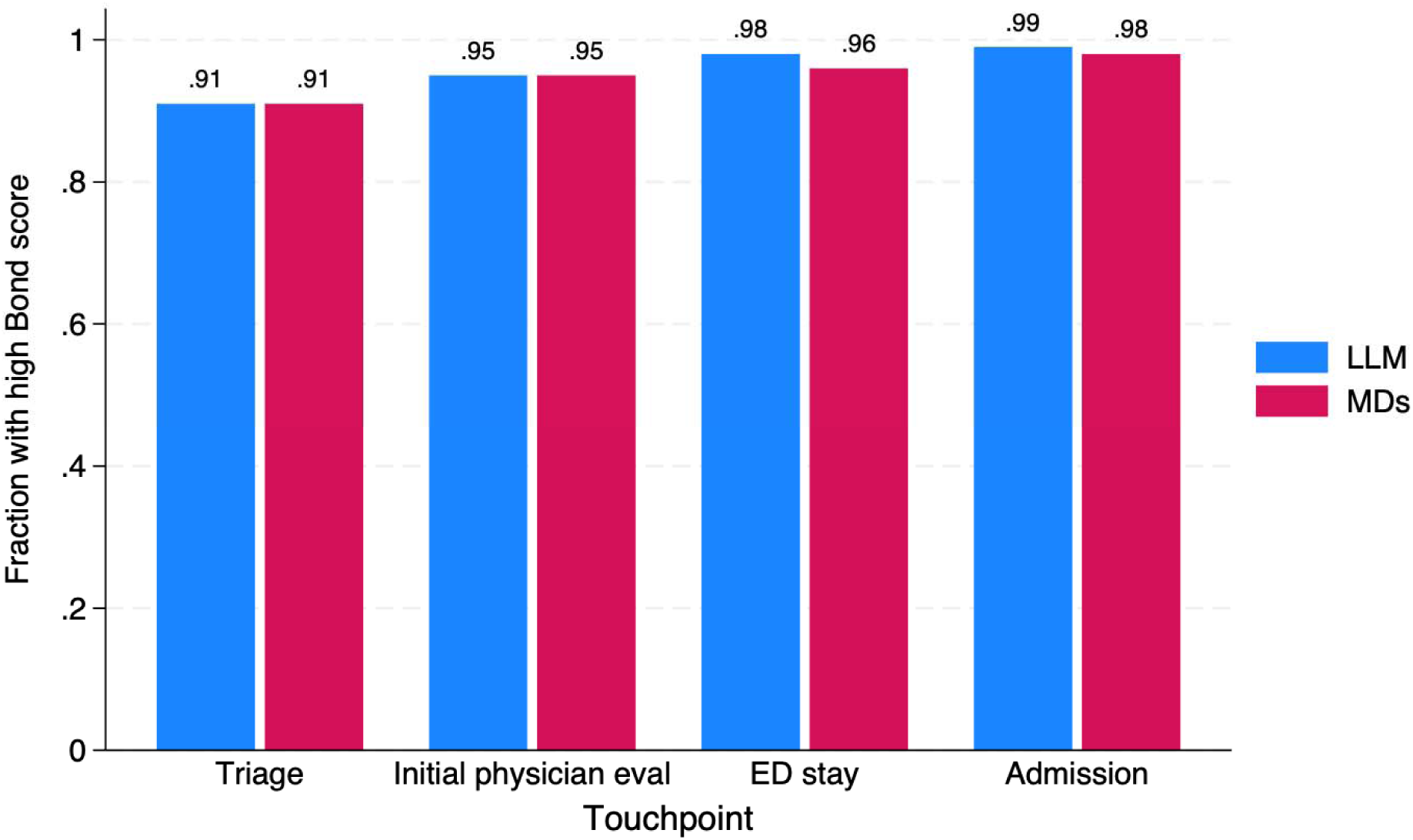
Proportion of differentials with high Bond scores (>= 4), by source: LLM (Dr. CaBot) vs MDs (pediatric emergency medicine physician diagnosticians).

### Rank of correct diagnosis

For the diagnosticians, the first-listed diagnosis was correct in 53, 61, 68, and 73 of the cases for touchpoints 1, 2, 3, and 4, respectively. For Dr. CaBot, the first-listed diagnosis was correct in 50, 62, 74, and 76 of the cases (**Figure 3**). There was no difference in pairwise comparisons (*P* > 0.15 for each touchpoint). These results are presented for each instance of Dr. CaBot in **Table S4** and for each diagnostician in **Table S5**.

**Figure 3.**
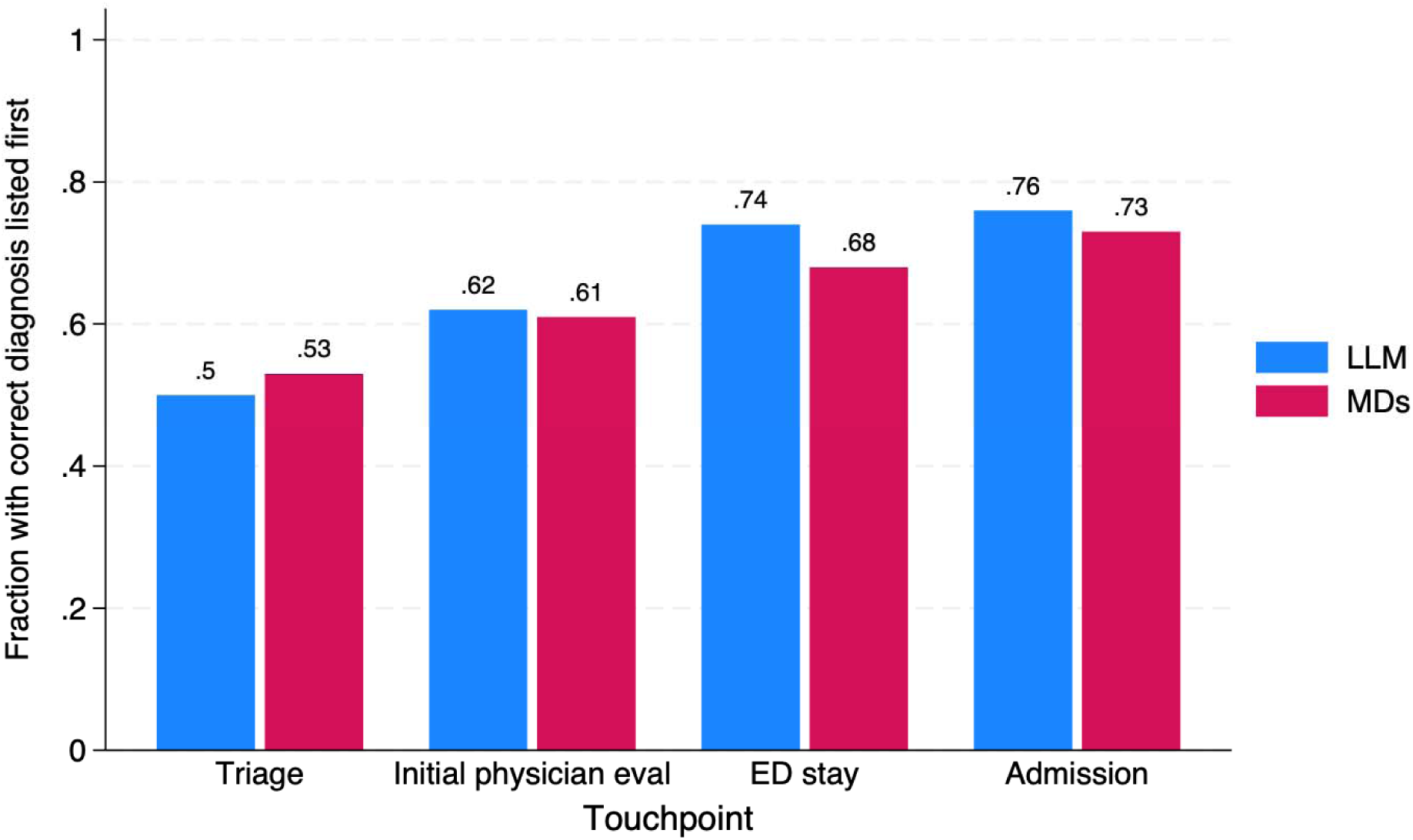
Proportion of differentials where the median rank of the correct diagnosis was first, by source: LLM (Dr. CaBot) vs MDs (pediatric emergency medicine physician diagnosticians).

## Discussion

For children presenting to the pediatric emergency department at a tertiary care urban children’s hospital, the agentic AI system Dr. CaBot and pediatric emergency medicine physicians showed similar performance in generating differential diagnoses. Accuracy was high for both sources from the first touchpoint. With triage information alone, each source had high-scoring differentials in 91% of cases, which left little room to detect a difference. The first-listed diagnosis was correct less often, in 50% to 76% of cases, and there too no difference was detected.

To our knowledge, only one prior study examined LLMs’ diagnostic reasoning on pediatric emergency cases,^16^ with two salient differences from the present study: 1) the prior study adapted real cases into vignettes, rather than using raw clinical notes; 2) the vignettes provided only information from initial evaluation (i.e. history and physical exam), and did not assess the LLMs’ ability to incorporate additional data such as laboratory results, imaging findings, or the physicians’ documented medical decision making.

This study has limitations. First, Dr. CaBot and the diagnosticians assessed clinical notes, not patients. Pediatric emergency specialists at a single center wrote the notes, sometimes after diagnostic evaluation had begun, and their reasoning was reflected in the documentation. Using notes from triage and the initial physician evaluation limited but did not eliminate this effect. Neither Dr. CaBot nor the diagnosticians could observe the child, hear the family’s account directly, or perform a physical examination. Performance with notes from general emergency departments remains untested. Second, we included only admitted patients, excluding discharged patients, among whom many delayed diagnoses occur.^10^ Third, masking was incomplete. Reviewers correctly identified the source of a differential 92% of the time, which may have biased Bond scores.

In conclusion, the Dr. CaBot AI agent generated highly accurate differential diagnoses for pediatric emergency patients based on clinical documentation, with performance similar to that of pediatric emergency medicine attendings. These findings suggest that Dr. CaBot could extend pediatric emergency specialist-level diagnostic reasoning to settings without it. However, it remains unclear how these accurate differentials could or should be integrated into patient care,^5,6^ and which patients and clinicians would most benefit from their use.

## Data Availability

The source clinical data cannot be shared externally due to the presence of protected health information. The anonymized, touchpoint-level Bond scores and diagnosis ranks are available upon reasonable request.

## Funding

NG: NIH T32HD040128.

KDM, AJMcM: Intelligent Health Laboratory Project, Boston Children’s Hospital.

TAB: Dunleavy Fund for Clinical AI at Harvard Medical School

AKM: NIH R01DK137993 and the Harvard Medical School Dean’s Innovation Awards for the Use of Artificial Intelligence in Education, Research, and Administration

## Declaration of interests

TAB: reports employment at Google LLC.

AKM: Visiting Researcher at Google DeepMind

## Declaration of generative AI and AI-assisted technologies in the manuscript preparation process

During the preparation of this work, the authors used Claude (Anthropic) for editing and revision under the authors’ direction and to draw Figure 1 from the authors’ original flowchart. The authors reviewed and edited the output as needed and take full responsibility for the content of the published article.

