## Supplemental Tables and Figures for "Agentic AI versus pediatric emergency physicians for differential diagnosis of acutely ill and injured children"

**Table S1**. Ground truth diagnosis organized by category of chief complaint.

*To be released at later date.*

**Table S2**. Comparison of the two configurations of Dr Cabot with and without literature search for the primary outcome of median Bond score >= 4, by touchpoint.

| **Touchpoint** | **Cabot with search** | **Cabot without search** |
| --- | --- | --- |
| Triage | 92/100 (92.0%) | 90/100 (90.0%) |
| Initial MD evaluation | 93/98 (94.9%) | 97/100 (97.0%) |
| ED stay | 98/100 (98.0%) | 97/100 (97.0%) |
| Admission | 97/98 (99.0%) | 99/100 (99.0%) |

**Table S3**. Comparison of the four diagnosticians individually for the primary outcome of median Bond score >= 4, by touchpoint.

| **Touchpoint** | **Diagnostician 1** | **Diagnostician 2** | **Diagnostician 3** | **Diagnostician 4** |
| --- | --- | --- | --- | --- |
| Triage | 83/98 (84.7%) | 88/99 (88.9%) | 86/100 (86.0%) | 89/99 (89.9%) |
| Initial MD  evaluation | 90/99 (90.9%) | 91/98 (92.9%) | 94/100 (94.0%) | 94/100 (94.0%) |
| ED stay | 93/99 (93.9%) | 93/99 (93.9%) | 95/100 (95.0%) | 97/100 (97.0%) |
| Admission | 95/99 (96.0%) | 93/99 (93.9%) | 95/99 (96.0%) | 97/100 (97.0%) |

**Table S4**. Comparison of the two configurations of Dr Cabot with and without literature search for the exploratory outcome of correct diagnosis, by touchpoint.

| **Touchpoint** | **Cabot with search** | **Cabot without search** |
| --- | --- | --- |
| Triage | 50/100 (50.0%) | 52/100 (52.0%) |
| Initial MD evaluation | 64/98 (65.3%) | 58/100 (58.0%) |
| ED stay | 71/100 (71.0%) | 70/100 (70.0%) |
| Admission | 75/98 (76.5%) | 76/100 (76.0%) |

**Table S5**. Comparison of the four diagnosticians individually for the exploratory outcome of correct diagnosis, by touchpoint.

| **Touchpoint** | **Diagnostician 1** | **Diagnostician 2** | **Diagnostician 3** | **Diagnostician 4** |
| --- | --- | --- | --- | --- |
| Triage | 45/98 (45.9%) | 50/99 (50.5%) | 44/100 (44.0%) | 55/99 (55.6%) |
| Initial MD  evaluation | 52/99 (52.5%) | 55/98 (56.1%) | 54/100 (54.0%) | 65/100 (65.0%) |
| ED stay | 58/99 (58.6%) | 66/99 (66.7%) | 64/100 (64.0%) | 70/100 (70.0%) |
| Admission | 60/99 (60.6%) | 69/99 (69.7%) | 70/99 (70.7%) | 77/100 (77.0%) |

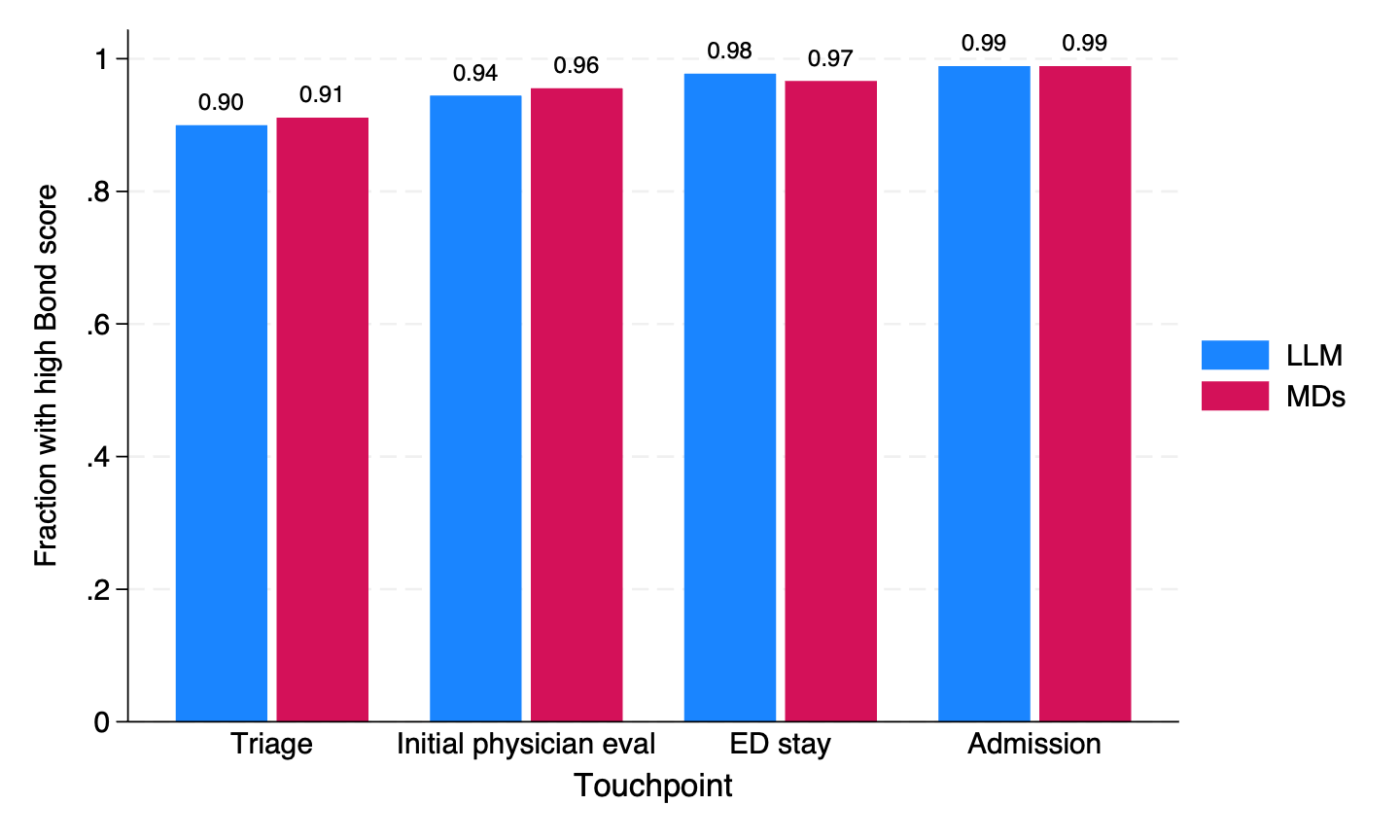

**Figure S1.** Sensitivity analysis excluding 10 transfers-in from another acute care hospital (N=90): proportion of differentials with high Bond scores (>= 4), by source: LLM (Dr CaBot) vs MDs (pediatric emergency medicine physician diagnosticians). No statistically significant differences between the LLM and MDs was detected by McNemar’s test (each *P*>0.5).
